# Detection of *KRAS, NRAS* and *BRAF* mutations in liquid biopsy from patients with colorectal cancer

**DOI:** 10.1101/2025.10.08.25337549

**Authors:** Katerina Ondraskova, Matous Cwik, Ondrej Horky, Jitka Berkovcova, Jitka Holcakova, Martin Bartosik, Tomas Kazda, Klara Mrazova, Michal Uher, Igor Kiss, Roman Hrstka

**Affiliations:** Research Centre for Applied Molecular Oncology (RECAMO), Masaryk Memorial Cancer Institute, Zluty kopec 7, 656 53 Brno, Czech Republic; National Centre for Biomolecular Research Faculty of Science, Masaryk University, Kamenice 5, 625 00 Brno, Czech Republic; Department of Comprehensive Cancer Care and Faculty of Medicine, Masaryk Memorial Cancer Institute and Masaryk University, Zluty kopec 7, 656 53 Brno, Czech Republic; Department of Pathology, Masaryk Memorial Cancer Institute, Zluty kopec 7, 656 53 Brno, Czech Republic; Department of Radiation Oncology, Masaryk Memorial Cancer Institute, Zluty kopec 7, 656 53 Brno, Czech Republic; Department of Biochemistry, Faculty of Science, Masaryk University, Kamenice 5, 625 00 Brno, Czech Republic; Department of Health Information, Masaryk Memorial Cancer Institute, Zluty kopec 7, 656 53 Brno, Czech Republic

**Author notes:** Correspondence: Assoc. Prof. Roman Hrstka, MSc., Ph.D.

**Keywords:** liquid biopsy, colorectal cancer, ddPCR, KRAS mutation

## Abstract

**Objectives:** Cancer treatment relies heavily on accurate diagnosis and effective monitoring of the disease. These processes often involve invasive procedures, such as colonoscopy, to detect malignant tissues followed by molecular analyses to determine relevant biomarkers. This study aimed to evaluate the clinical performance of droplet digital PCR (ddPCR) for detecting *KRAS, NRAS*, and *BRAF* mutations in circulating tumor DNA (ctDNA) from colorectal cancer patients using liquid biopsy.

**Methods:** ctDNA was isolated from CRC patients (n = 110) and analyzed for *KRAS, BRAF*, and *NRAS* mutations. The ctDNA obtained through liquid biopsy was analyzed using ddPCR, and the findings were compared with sequencing data from tumor DNA archived in formalin fixed paraffin-embedded blocks (FFPE).

**Results:** For *KRAS* mutations, ddPCR achieved a sensitivity of 72% and a specificity of 71.4%. However, when pooling all target mutations (*KRAS, NRAS* and *BRAF*), the overall sensitivity and specificity were lower, at 48.3% and 51.1%, respectively.

**Conclusion:** The results of this study indicate that the ddPCR analysis of ctDNA may provide complementary information for the molecular diagnosis of CRC patients.

## 1. Introduction

Accurate diagnosis is the basis of cancer treatment, beginning with the identification of malignant tissue present in a patient [1]. The usual way to confirm the presence and characteristics of cancer is through tissue biopsy analysis, where tumor cells exhibit distinct properties from healthy cells. These differences arise due to genetic changes conferring a selective advantage and accumulating throughout tumor progression. These mutations may serve as important predictive and prognostic factors in oncological practice [2]. However, traditional tissue biopsy procedures are invasive and may capture only a limited and non-representative fraction of tumor cells. Moreover, excised tumor tissues do not reflect the dynamic nature of the tumor microenvironment and may differ significantly from residual tumor tissue, which can serve as a source of recurrence. This has led to research on other potential ways for cancer detection – one of them being analysis of liquid biopsies (LBs) [3].

Liquid biopsy offers a non-invasive technique to analyze body fluids (most often blood or urine), including components such as various protein biomarkers, circulating cell-free DNA (cfDNA), circulating tumor DNA (ctDNA), circulating tumor cells (CTCs) and many others [4]. LBs allow monitoring of patients with cancer, especially based on DNA or RNA analysis, and represent an alternative when tissue biopsies cannot be obtained or are complicated, such as colonoscopy. Circulating tumor DNA, a fraction of cell-free DNA originating specifically from tumor cells, provides an innovative approach for cancer monitoring [5]. Specifically, these small DNA fragments with tumor-specific mutations are released into the blood and constitute only a minor part of the total cfDNA. According to recent studies, ctDNA accounts for less than 1% of an individual’s total cfDNA. Recent advancements in assay sensitivity and refinement have established ctDNA analysis as a reliable tool in the field of oncology [6]. Therefore, ctDNA can be used as a prognostic and diagnostic biomarker due to the correlation between the stage of cancer and other factors, such as location, tumor progression, and pattern of metastasis [7, 8]. In the last decade, there has been a significant improvement in the analysis of cfDNA and, consequently, ctDNA. Next-generation sequencing (NGS) and digital PCR (dPCR) are some of the most frequently used methods for detecting cfDNA and ctDNA in liquid biopsy, mainly due to the high specificity and sensitivity [9]. Among these, droplet digital PCR (ddPCR) offers additional advantages, such as absolute quantification, high precision, and the ability to detect low-frequency mutations, making it especially valuable for monitoring minimal residual disease and treatment response.

Colorectal cancer (CRC) is one of the most frequently diagnosed malignancies worldwide and currently ranks third in terms of incidence. In addition, CRC is the second leading cause of cancer-related mortality worldwide. In 2022, according to the GLOBOCAN database, more than 1.9 million cases of CRC were recorded. There is a stable rise in the incidence of CRC among adults younger than 50 years, and it has been rising in developing countries as well. A substantial number of patients (approximately 25%) are still diagnosed with metastatic disease and an additional 25-50% of patients with early stage disease will develop metastases later [10-12]. The treatment and prognosis of metastatic CRC remain difficult, with a 5-year relative survival rate of only 12.5% in the US [10]. Thus, there is a need to improve screening, diagnostics, and treatment. Standard screening and/or diagnostic tools include fecal occult blood tests (FOBTs), colonoscopy or CT scan.

Current metastatic CRC treatment involves biomarker testing, including *RAS* mutations [13], *BRAF*^V600E^ mutations and microsatellite instability (MSI) status [14]. The presence of *RAS* mutations predicts a lack of response to anti-EGFR treatments, while *BRAF*^V600E^ mutation is a negative prognostic marker. However, it is a positive predictive marker for novel *BRAF*^V600E^ – targeted therapies [15]. MSI status has prognostic and predictive value for responsiveness to modern immunotherapy with checkpoint inhibitors. *KRAS* (*Kirsten rat sarcoma*) is a gene located on chromosome 12 that provides signals allowing the formation of the K-Ras protein. Activated *KRAS* mutations drive cell proliferation, suppress differentiation and are crucial for the initiation of carcinogenesis. K-Ras, a membrane-anchored GTP/GDP-binding protein, is responsible for the transmission of signals from the cytosol to the cell nucleus. It is also part of the important MAPK/ERK signaling pathway [13, 14, 16, 17]. *NRAS* (*Neuroblastoma RAS Viral Oncogene Homolog*) mutations are associated with tumor development and protection of cells from apoptosis [18]. They occur in only 3-5% of all CRCs, making them less frequent than *KRAS* or *BRAF* mutations, with which they are also believed to be incompatible [19]. However, the importance of *NRAS* in CRC has increased owing to its possible role in the clinical management of advanced stages of this disease [20].

The effect of CRC treatment seems to be closely related to the mutation status of the *BRAF* (*B-Raf Proto-Oncogene Serine/Threonine-Protein Kinase*) gene and MSI status. CRC patients with MSI-stable carcinoma and the presence of mutation in the *BRAF* gene show shorter overall survival (OS) than patients without the mutated gene [21, 22]. In addition to *KRAS* mutations, *BRAF* mutations are also recognized as critical factors influencing the efficacy of anti-EGFR targeted therapy, being responsible for worse prognosis and reduced response to chemotherapeutic agents [23].

This retrospective study evaluates the diagnostic potential of cfDNA obtained from liquid biopsy using ddPCR in the molecular diagnosis of CRC to determine the benefit of ctDNA-based diagnosis compared to standard tissue biopsy. We determined the sensitivity, specificity, and overall efficiency of ddPCR for the detection of the most common *KRAS* gene point mutations at the major hotspot codons G12 and G13. We also identified mutations in *BRAF* and *NRAS*, which are frequently present in CRC, especially in cases with wt *KRAS*. We optimized the method using selected cancer cell lines containing mutations in these genes (more information is provided in the Methods section). Following this, we tested samples from patients with CRC to explore its applicability in molecular diagnostics, monitoring of minimal residual disease, therapeutic decision-making, and ultimately the quality of life of patients in the future.

## 2. Materials and Methods

### Cell lines

The cell lines used to optimize the detection of selected mutations by ddPCR (Table S1) were obtained from the American Type Culture Collection (ATCC). They were maintained in medium as indicated (all media were from Merck & Co., Inc., Rathway, New Jersey, U.S.), supplemented with 10% fetal bovine serum (Life Technologies Corp., Carlsbad, California, U.S.), 1% pyruvate and L-glutamine at 37 °C in a humidified atmosphere of 5% CO_2_. Subcultures were prepared when cells reached 75% confluence, roughly every 2-3 days. All cell lines were regularly tested for mycoplasma contamination with consistently negative results, and those obtained more than two years ago additionally underwent authentication by short tandem repeat (STR) profiling. These cell lines served as positive controls to validate the ddPCR assays and to ensure assay specificity and accuracy.

### Genomic DNA extraction and fragmentation

Genomic DNA (gDNA) was isolated from individual cell lines using the DNeasy Blood & Tissue Kit (QIAGEN N.V., Venlo, Netherlands). The concentration of the eluted gDNA was measured using a Qubit™ 4 Fluorometer (Thermo Fisher Scientific, Waltham, Massachusetts, U.S.) and Qubit™ dsDNA BR Assay Kit (Thermo Fisher Scientific, Waltham, Massachusetts, U.S.). gDNA was then diluted to achieve a concentration of 2 µg×µl^-1^ and subjected to the fragmentation using a high-capacity ultrasonicator Covaris M220 (Covaris, LLC, Woburn, Massachusetts, U.S.). The process was recorded using a SonoLab7 system (Covaris, LLC, Woburn, Massachusetts, U.S.), with Snap-Cap micro-TUBE tubes (Sigma-Aldrich, Burlington, Massachusetts, U.S.). There were 200 cycles during the fragmentation of each sample. The fragmentation efficacy was assessed using agarose gel electrophoresis (data not shown).

### Liquid biopsies

Venous blood samples from CRC patients (Table 1) were collected between 2011 and 2020 in Sarstedt tubes containing a clotting activator, allowed to clot at room temperature for 45 minutes, and subsequently centrifuged to obtain serum. The extracted serum was immediately stored at -20°C and moved to -80°C within two weeks. The samples were then stored in the Bank of Biological Material at Masaryk Memorial Cancer Institute (BBM MMCI) at -80 °C. Out of total 110 patients, 39 were women with a median age of 64.5 years and 71 were men with a median age of 66.0 years. An addi-tional cohort of 30 control samples (40% female, 60% male) was collected from volunteers who underwent preventive cancer screening at MMCI to test the specificity of ddPCR. All participants enrolled in the study signed an informed consent to provide samples for research purposes and the study was approved by the Institutional Review Board (2020/2489/MOU). To minimize degradation and freeze–thaw artifacts, all serum aliquots were subjected to a single freeze-thaw cycle prior to cfDNA extraction. Hemolytic samples were excluded based on the visual inspection. cfDNA was isolated using the Plasma/Serum Cell-Free Circulating DNA Purification Mini Kit (Norgen Biotek Corp., Thorold, Ontario, Canada) and its concentration was measured using the Qubit™ dsDNA HS Assay Kit (Thermo Fisher Scientific, Waltham, Massachusetts, U.S.), which also served as a basic cfDNA integrity metric. The concentrations of isolated cfDNA ranged from 0.08 to 33.90 ng×µl^-1^. All pre-analytical processes were carried out in accordance with the general guidelines for the handling of cellfree nucleic acids, including the ISO 20186-3:2019 recommendations. Recent evidence emphasizes the critical importance of standardized pre-analytical workflows in minimizing cfDNA degradation and pre-analytical variability, particularly regarding serum vs. plasma handling, centrifugation timing, and temperature control [24].

**Table 1.** Patient medical data.

| Basic characteristics of patients and primary tumor<br>(n = 110 patients) |  | Descriptive statistics* |
| --- | --- | --- |
| **Sex** | women | 39 (35.5 %) |
|  | men | 71 (64.5 %) |
| **Age at surgery** | [years] | 64.5 ± 10.7 |
|  |  | 66.0 (58.2; 73.0) |
| **Localization of the primary tumor** | Cecum (C18.0, C18.1) | 20 (18.2 %) |
|  | colon (C18.2–C18.7, C18.9) | 28 (25.5 %) |
|  | lesions extending beyond the colon (C18.8) | 8 (7.3 %) |
|  | rectosigmoidal connection (C19) | 22 (20.0 %) |
|  | rectum (C20) | 32 (29.1 %) |
| **Extent of tumor (T)** | T1 | 1 (0.9 %) |
|  | T2 | 8 (7.3 %) |
|  | T3 | 72 (65.5 %) |
|  | T4 | 27 (24.5 %) |
|  | Tx | 2 (1.8 %) |
| **Metastases in the lymph nodes (N)** | N0 | 37 (33.6 %) |
|  | N1 | 34 (30.9 %) |
|  | N2 | 37 (33.6 %) |
|  | Nx | 2 (1.8 %) |
| **Distant metastases (M)** | M0 | 57 (51.8 %) |
|  | M1 | 47 (42.7 %) |
|  | Mx | 6 (5.5 %) |
| **Lymphovascular space invasion (L)** | L0 | 62 (56.4 %) |
|  | L1 | 35 (31.8 %) |
|  | Lx | 13 (11.8 %) |
| **Grade** | G1 | 27 (24.5 %) |
|  | G2 | 60 (54.5 %) |
|  | G3 | 18 (16.4 %) |
|  | G4 | 1 (0.9 %) |
|  | Gx | 4 (3.6 %) |
| **Clinical stage** | stage I | 3 (2.7 %) |
|  | stage II | 24 (21.8 %) |
|  | stage III | 36 (32.7 %) |
|  | stage IV | 47 (42.7 %) |
| **Neoadjuvant therapy** | no | 93 (84.5 %) |
|  | yes | 17 (15.5 %) |

\* Mean ± standard deviation and median (interquartile range) are given for continuous variables (age) and absolute (relative) frequencies are given for categorical variables.

### NGS analysis

The percentage of neoplastic cells in formalin-fixed paraffin-embedded (FFPE) tissues was estimated by a pathologist, and the minimum content of tumor cells in the selected area required for molecular analysis was 10%. DNA was extracted using the cobas^®^ DNA Sample Preparation Kit (Roche) or the QIAamp DNA FFPE Tissue Kit (QIAGEN N.V., Venlo, Netherlands).

Mutational analysis was performed by NGS using commercially available kits according to manufacturer’s protocols: EliGene Colorectum NGS (Elisabeth Pharmacon, Brno, Czech Republic) or Ac-cel-Amplicon (Swift Biosciences, Ann Arbor, Michigan, U.S.) as amplicon sequencing approach, or hybridization-based Sequence Capture targeted enrichment of custom-defined regions (Roche NimbleGen, Inc., Pleasanton, California, U.S.). Sequencing was performed using a MiSeq instrument (Illumina, San Diego, California, U.S.). The mutation status was assessed using NextGENe software (Softgenetics LLC, State College, Pennsylvania, U.S.). The minimum allele frequency was set to 5% and the minimum coverage of sequenced regions (300×) was checked in IGV (Broad Institute) and/or NextGENe. All three NGS panels covered the hotspot regions subsequently assessed by ddPCR (KRAS codons 12/13, NRAS codon 61, and BRAF V600E). No systematic differences in mutation detection were observed between the sequencing platforms, and NGS results across platforms were deemed comparable reference standards.

### Droplet digital PCR (ddPCR)

We focused on detecting the most frequent and clinically relevant hotspot mutations in *KRAS* (codons 12 and 13), *NRAS* (codon 61), and *BRAF* V600E, as they represent the majority of actionable mutations associated with anti-EGFR therapy resistance in colorectal cancer. Other *KRAS* codons and non-V600E *BRAF* mutations, although biologically relevant, were not included due to their low prevalence (<5%) and limited impact on treatment algorithms as supported by [22, 23]. For the analysis, we used commercial kits produced by Bio-Rad: the ddPCR™ KRAS G12/G13 Screening Kit (1863506), ddPCR™ BRAF V600 Screening Kit (12001037), and ddPCR™ NRAS Q61 Screening Kit (12001006). The method was optimized and validated using cancer-derived cell lines differing in *KRAS, BRAF* or *NRAS* gene mutation statuses (Table S1), which served as positive and negative controls for each assay. These reference cell lines were used to define the droplet amplitude thresholds and confirm the accuracy of the mutation detection. Droplet clustering patterns were confirmed in each run using these controls, as illustrated in Figure 1.

**Figure 1.**
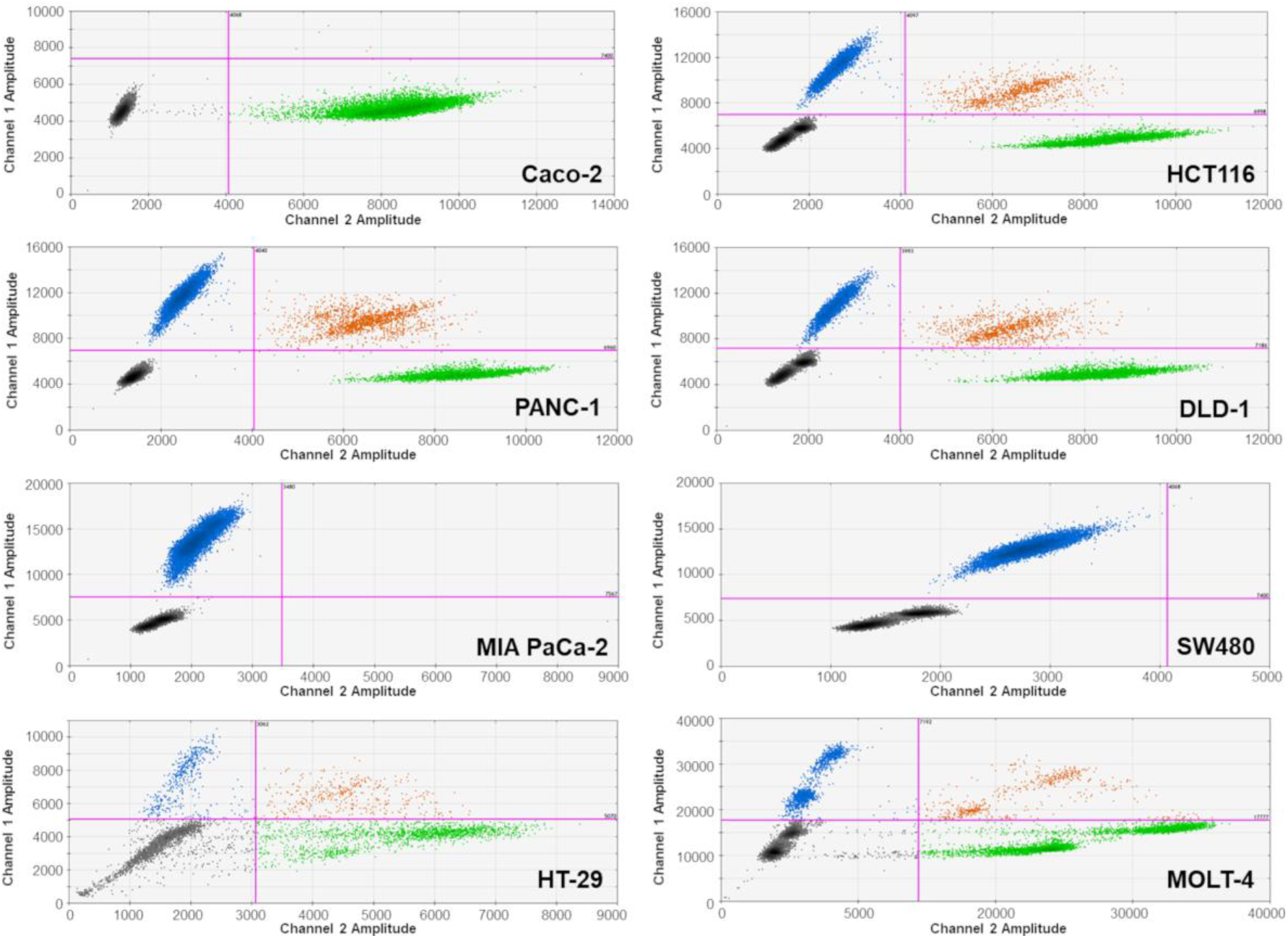
Representative ddPCR graphs of genomic DNA (gDNA) from tested cancer cell lines. Droplet populations are color-coded as follows: blue – droplets containing mutant alleles; orange – droplets containing both mutant and wild-type alleles; green – droplets containing only wild-type alleles; black – negative droplets (no target DNA). Droplet clusters were manually gated using threshold lines (purple). Mutation-specific assays were applied as follows: KRAS hotspot muta-tions (codons G12/G13) were analyzed in all cell lines except HT-29 and MOLT-4; BRAF V600E mutation was analyzed in the HT-29 cell line; and NRAS G12C mutation in the MOLT-4 cell line.

The reaction mix contained 11 µL of 2× ddPCR Supermix for Probes (No dUTP), 1 µL of 20× Multiplex Primer FAM + HEX (all Bio-Rad Laboratories, Inc., Hercules, California, U.S.), and 4.8 – 5 µL of deionized water depending on whether 0.2 µL of restriction enzyme *MseI* (*KRAS* mutation, New England Biolabs, Ipswich, Massachusetts, U.S..) or *HindIII* (*NRAS* and *BRAF* mutations, MBI Fermentas, Amherst, New York, U.S.) were added to the mix in the case of genomic DNA. The mixture was then pipetted into a High-Profile 96-Well PCR plate (Bio-Rad Laboratories, Inc., Hercules, California, U.S.) and 5 µL of cfDNA were added to a final volume of 22 µL per well. Subsequently, the plate was sealed by PX1™ PCR Plate Sealer (Bio-Rad Laboratories, Inc., Hercules, California, U.S.) and inserted into an Automated Droplet Generator system (Bio-Rad Laboratories, Inc., Hercules, California, U.S.) to prepare a water-oil emulsion of the samples. PCR was performed in C1000 Touch™ Thermal Cycler according to the manufacturer’s protocol (Table S2). The plate was then inserted into the QX200 Droplet Reader device to analyze all droplets using a two-color detection system. The results were plotted on a graph of fluorescence intensity using QuantaSoft™ Software (all Bio-Rad Laboratories, Inc., Hercules, California, U.S.). Droplet amplitude thresholds were manually defined with reference to the positive and negative controls for each run (Figure 1). The threshold line was set above the highest cluster of negative droplets and below the lowest cluster of positive droplets, in accordance with the Bio-Rad guidelines.

To ensure that the performance of the measurement procedure is consistent with detection capability we determined both the Limit of Blanc (LoB) and the Limit of Detection (LoD) for the ddPCR assay. The LoB was established by performing 20 measurements using gDNA (wt *KRAS* gene), with 18 out of 20 results falling below the LoB threshold of 0.4 copies/μl. This corresponds to a 90% proportion with an associated upper one-sided 95% confidence interval of 96.6%.

To define the sensitivity of detection, we conducted serial dilutions of genomic DNA derived from the DLD-1 cell line (KRAS G13D mutation), as summarized in Table S3. Three independent dilution experiments were performed in three technical replicates each. The minimal detectable input of cfDNA in the ddPCR reaction was determined to be approximately 1.76 ng/μl. A minimum of two positive droplets (i.e. ≥ 0.4 copies/μl) was required to classify a sample as mutation-positive. These empirically derived thresholds were used to interpret patient samples and ensured that only signals above the assay-specific detection limit were considered true positives.

To assess inter-operator reproducibility, two independent analysts reviewed 110 ddPCR results. Operator 1 identified 67 samples as mutant and 43 as wild-type. Operator 2 classified 66 of the 67 mutant samples identically and all 43 wild-type samples as such, with only one discordant call. Cohen’s κ statistic was 0.981, confirming excellent agreement and high robustness of the classification process. A cell line displaying the tested mutation (Table S1, Figure 1) was used as a positive control for each cfDNA analysis. Deionized water was used as a negative control.

### Statistical analysis

Standard descriptive statistics were used to describe the analyzed cohort: mean, standard deviation, median and interquartile range are given for continuous variables and absolute and relative frequencies are given for categorical variables. The diagnostic accuracy of mutation detection by ddPCR was evaluated using sensitivity, specificity, predictive values and, using ROC analysis, the overall accuracy was summarized and tested as the area under the curve (AUC). A univariate logistic regression model was used for the basic assessment of the association of clinical and pathological features with mutational status, while a multivariate model was used to estimate odds ratios (OR) adjusted for other variables. Statistical significance testing of the AUC and ORs were performed at the 5% significance level. All calculations were performed in IBM SPSS Statistics software version 25.

## 3. Results

### Optimization of ddPCR

To assess the sensitivity of ddPCR (i.e., to what extent the *KRAS* mutation can still be detected by this method), dilution series of genomic DNA (gDNA) were prepared. The limit of detection (LoD) was determined using isolated gDNA from DLD-1 (heterozygous KRAS G13D) cell line. Initial input amount was 400 ng of DNA/μL (100%, final concentration of 11.429 ng/μl), and subsequent dilutions were prepared to final concentrations of 25%, 6.25%, 1.56%, 0.39%, 0.098% and 0.024% (Table S3). ddPCR analysis confirmed that positive and negative droplets could still be clearly distinguished even at low gDNA input levels, demonstrating high assay sensitivity. Based on the dilution series, our results indicate that the minimal reliably detectable concentration is approximately 0.04 ng/µL. Subsequently, we analyzed a panel of cell lines with known mutation status (mutant, wild-type, or heterozygous; Table S1) in order to verify the assay’s selectivity and to define the amplitude ranges for both wild-type and mutant droplets. Considering these findings and the determined Limit of Blank, a threshold value of ≥ 0.04 copies/µL was established to classify samples as positive for mutation detection.

### Samples from patients with CRC

A total of 110 patients with primary colorectal cancer were included in this study, of whom 47 patients (42.7%) had metastatic disease. Patients were treated according to the standard of care. All relevant clinical, demographic, and epidemiological data were collected from each patient (Table 1). As a control group, we used healthy patients participating in a cancer prevention program at the Masaryk Memorial Cancer Institute (30 patients) who did not have cancer or any severe health problems. Mutational status of *KRAS/BRAF/NRAS* genes in the primary tumor was determined by NGS, applying either an amplicon or hybridization-based strategy, and the results were compared with the mutational status identified using ctDNA analysis by ddPCR from the peripheral blood serum of the respective patients (Table 2). No systematic differences in mutation detection were observed between the platforms, and all provided concordant results with the ddPCR assays. Briefly, 63 tumor samples were mutated (57.3%) in at least one of the analyzed genes, and 47 samples were wild-type (42.7%) using the standard of care predictive pathology tissue examination *via* NGS. For comparison, analysis of ctDNA from liquid biopsies revealed that 68 samples were mutated (61.8%) and 42 samples were wild-type (38.2%). The most abundant mutation in both cases was the *KRAS* gene mutation, which was present in 50 samples by NGS and 67 samples by ddPCR (45.5%; 60.9%). Other mutations were present in the following order: *BRAF*^V600E^ mutation was present in 9 and 5 samples respectively (8.2%; 4.5%), and *NRAS* mutation was found in 4 and 3 samples respectively (3.6%; 2.7%). One sample showed concomitant mutations detected by ctDNA analysis. All liquid biopsy samples were primarily tested for the presence of *KRAS* mutation. Majority of samples with confirmed *KRAS* mutation were not further tested for the presence of *BRAF* and *NRAS* mutations (Table S4), as the primary aim of the study was to determine any mutation and use it for subsequent disease monitoring.

**Table 2.** Determination of *KRAS, BRAF* and *NRAS* mutations.

| Gene |  | NGS | ddPCR |
| --- | --- | --- | --- |
| ***KRAS*** | mut | 50 (45.5%) | 67 (60.9%) |
|  | wt | 60 (54.5%) | 43 (39.1%) |
|  | NA | 0 (0.0%) | 0 (0.0%) |
| ***BRAF*** | mut | 9 (8.2%) | 5 (4.5%) |
|  | wt | 99 (90.0%) | 28 (25.5%) |
|  | NA | 2 (0.9%) | 77 (70%) |
| ***NRAS*** | mut | 4 (3.6%) | 3 (2.7%) |
|  | wt | 105 (95.5%) | 10 (9.1%) |
|  | NA | 1 (0.9%) | 97 (88.2%) |
| ***KRAS/BRAF/NRAS*** | mut | 63 (57.3%) | 68 (61.8%) |
|  | wt | 47 (42.7%) | 42 (38.2%) |
|  | NA | 0 (0.0%) | 0 (0.0%) |

The absolute (relative) frequencies of the test results are provided, including the unperformed tests (especially for the ddPCR method).

In several cases, mutations were detected in ctDNA but were absent in tumor tissue (seventeen patients, Table 2). Conversely, there were also patients in whom mutations were present in the tumor tissue but not detectable in ctDNA. Importantly, no mutations were detected in the control group. Previous studies have also demonstrated that disagreement between tissue and plasma-based detection of KRAS mutations may occur and may influence treatment decisions. For example, the IRON group reported an 85% concordance (29/34 patients) using chip-based digital PCR, with sensitivity and specificity of 69% and 100%, respectively [25]. Similarly, the PREDATOR study detected KRAS mutations in plasma not always observed in tissue, likely reflecting tumor heterogeneity or clonal evolution [26]. Several other studies confirm discordance rates of 10–20% in mCRC cohorts, often due to low ctDNA burden or sampling bias [27, 28]. These findings highlight the need for orthogonal validation, such as ddPCR for hotspot regions, which we employed to ensure consistency across NGS platforms.

A detailed comparison of concordant and discordant cases is provided in Supplementary Tables S5 and S6, which summarize the tumor stage, neoadjuvant treatment status, and tissue–blood sampling interval. Discordant results occurred more frequently among patients who received neoadjuvant therapy and those with longer intervals between blood and tissue sampling. The ddPCR method achieved a statistically significant but only modest discrimination performance when detecting KRAS mutations, with an AUC = 0.602 (p < 0.026), sensitivity of 72.0%, and specificity of 48.3% (Table 3). When all three target mutations were included in the analysis, the overall accuracy improved slightly (AUC = 0.612; p = 0.016) accompanied by comparable sensitivity (71.4%) and a minor increase in specificity (51.1%) (Table 3). These results indicate that while ddPCR can detect a substantial fraction of mutation-positive cases, its ability to correctly exclude mutation-negative patients is limited. The subgroup analyses stratified by clinical stage (I–IV) and mutation class are summarized in Supplementary Table S7, showing improved sensitivity in stage IV disease but persistently low specificity across all subgroups. Briefly, in early stage disease (stage I–II), the diagnostic performance was modest, with a sensitivity of 66.7% and a specificity of 55.6%. In stage III, the accuracy remained limited, with sensitivities of approximately 55–58% and specificities of 50–58%, depending on whether only *KRAS* or any *KRAS*/*BRAF*/*NRAS* mutation was considered. In contrast, the performance improved in advanced disease: in stage IV, the sensitivity increased to 87.0% for *KRAS* mutations and 83.3% for *KRAS*/*BRAF*/*NRAS*, albeit at the cost of reduced specificity (∼41%). In the subgroup of patients without neoadjuvant therapy (n=93), the sensitivity reached 78.6% for *KRAS* mutations, with a specificity of 52.9%.

**Table 3.** Evaluation of the diagnostic accuracy of *KRAS* or any *KRAS/BRAF/NRAS* mutation detection by ddPCR.

| Metric | *KRAS* mutation | *KRAS/BRAF/NRAS* mutation |
| --- | --- | --- |
| Area under the curve (AUC) | 0.602 (0.691; 0.512) | 0.612 (0.521; 0.704) |
| AUC with bootstrapped CI | 0.602 (0.513; 0.690) | 0.612 (0.522; 0.703) |
| Sensitivity | 72.0 (58.1; 82.7) % | 71.4 (59.1; 81.2) % |
| Specificity | 48.3 (36.1; 60.8) % | 51.1 (37.1; 64.9) % |
| Positive predictive value | 53.7 (41.8; 65.2) % | 66.2 (54.2; 76.4) % |
| Negative predictive value | 67.4 (52.3; 79.7) % | 57.1 (42.0; 71.1) % |

For all values 95% CI is given.

### Association of clinical and pathological features with the mutational status

We evaluated which clinicopathological factors were significantly associated with the presence of the *KRAS* gene mutation (Supplementary Table S8). *KRAS* mutations were identified with greater frequency in rectal (36.0% vs. 23.3%) and cecal tumors (22.0% vs. 15.0%), whereas tumors at the rectosigmoid junction were more prevalent in the *wild-type* group (28.3% vs. 10.0%). However, these differences were not statistically significant. Other relevant findings included a higher proportion of N2 lymph node metastases among patients in the *KRAS-mutated* group (42.0 vs. 26.7%) and a trend towards a higher prevalence of clinical stage IV cases (46.0% vs. 40%). Although not statistically significant, these observations may suggest increased tumor aggressiveness associated with the presence of the *KRAS* mutation [29].

When extending the analysis to include any of the *KRAS*/*BRAF*/*NRAS* mutations (Supplementary Table S9), the association with cecal tumor localization became statistically significant (25.4% vs. 8.5%, p = 0.009). Moreover, patients harboring any of these mutations were more often diagnosed at clinical stage IV (47.6% vs. 36.2%, p = 0.017) and again showed a trend towards a higher prevalence of N2 lymph node metastases (41.3% vs. 23.4%, p = 0.049). Taken together, the presence of *KRAS/BRAF/NRAS* mutations was associated with more aggressive disease features, particularly in cecal tumors, consistent with recent evidence linking mutation status to tumor localization and nodal involvement [30].

## 4. Discussion

Circulating nucleic acids represent a promising new type of biomarker with the potential to improve cancer treatment in many aspects in the future. One of the aims of this study was to evaluate the use of ddPCR in the detection of somatic mutations present in the tumor tissue of CRC patients. Tissue-based mutation analysis is limited by certain characteristics of cancer, particularly the spatial and temporal heterogeneity of tumor tissue [29]. Moreover, for patients where tumor tissue sample is difficult or impossible to obtain, ctDNA-based testing might provide a non-invasive solution for the purpose of initial molecular diagnosis. The percentage of CRC patients in whom ctDNA can be detected depends on the extent of the disease and ranges from 50% in patients with non-metastatic disease to nearly 90% in patients with metastatic disease [31]. Therefore, ctDNA analysis could have applications in screening, molecular diagnosis of cancer, risk stratification of disease relapse after initial curative intent treatment or monitoring of cancer treatment success in a palliative setting [32]. Evidence from multiple trials indicates a high consistency (> 90%) between ctDNA and standard tumor tissue-based *RAS* testing [8].

Among the available technologies for ctDNA analysis, ddPCR has emerged a sensitive and specific method for detecting tumor-associated mutations [32]. However, its use in clinical practice remains limited [33]. This makes liquid biopsy and ddPCR-based ctDNA analysis an increasingly attractive complementary approach, particularly as a faster alternative for patients requiring urgent treatment decisions or those whose molecular profile restricts therapeutic options. Beyond its non-invasive nature, liquid biopsy also enables longitudinal monitoring of tumor evolution and better captures the heterogeneity of primary tumors and metastatic lesions [29]. Building on this rationale, our results demonstrate that ddPCR is a robust and sensitive method for detecting *KRAS* mutations in ctDNA, although its specificity was moderate. The slight improvement in overall accuracy when combining multiple mutations suggests that expanding the mutation panel may help to partially compensate for the limited specificity inherent to ctDNA analysis in early-stage disease or low-tumor-burden settings. Although the diagnostic gain was incremental, these findings reinforce the utility of ddPCR not only in mutation screening but also in tracking minimal residual disease over time, where even small changes in ctDNA levels may be clinically relevant.

Despite achieving statistically significant discrimination, the diagnostic accuracy of ddPCR in our study was modest, with AUC values close to 0.6 and specificity consistently below 52%. These findings underscore important limitations of ddPCR as a stand-alone diagnostic tool in this setting. Several technical and biological factors likely contributed to the observed performance. First, serum rather than plasma was used for cfDNA isolation, and serum-derived samples are known to suffer from leukocyte DNA contamination that reduces assay specificity [34]. Second, a substantial proportion of patients had early-stage disease, where ctDNA shedding is often insufficient for reliable detection, as also shown by Bettegowda et al., who detected ctDNA in only 55% of localized cases [6]. Low ctDNA levels, together with limited assay sensitivity, likely explain cases where KRAS mutations were detected in tissue but not in ctDNA. Conversely, instances where mutations were found in cfDNA but not in tumor tissue may reflect tumor heterogeneity or spatial sampling limitations [35]. Biological variability introduced by neoadjuvant therapy and longer blood–tissue sampling intervals further contributed to discordant results, as effective treatment can reduce tumor burden and ctDNA levels to below detection thresholds [36, 37]. Our subgroup analyses (Supplementary Tables S5 and S6) support this interpretation, showing that discordant findings were more frequent in patients who had undergone neoadjuvant therapy and in those with longer sampling intervals.

In addition, certain methodological aspects may have influenced the results. The mutation panel was restricted to the hotspots covered by the selected Bio-Rad kits, precluding analysis of less frequent KRAS or BRAF variants. In addition, not all patients underwent ddPCR testing for *BRAF* and *NRAS* mutations. This was an intentional choice, as once a *KRAS* mutation was identified, further *NRAS*/*BRAF* testing was not performed in line with current diagnostic algorithms that consider these mutations largely mutually exclusive. While rare instances of concurrent mutations have been reported [38], their frequency remains low, and universal multi-gene testing in *KRAS-*positive cases is not yet routine.

Finally, clonal hematopoiesis of indeterminate potential may represent another important confounder. Somatic variants arising in hematopoietic stem cells can be released into circulation and misclassified as tumor-derived alterations [39, 40]. Somatic mutations in hematopoietic stem cells, most frequently involve genes such as *DNMT3A, TET2*, and *ASXL1*, but occasionally also affect genes involved in RAS/MAPK pathways. These mutations can be released into circulation and detected in cfDNA, potentially mimicking tumor-derived alterations. Since our study did not include matched sequencing of leukocyte DNA, we cannot exclude the possibility that some of the mutations detected in serum samples originated from clonal hematopoiesis. Recent studies have shown that clonal hematopoiesis is common in older cancer patients and can confound liquid biopsy analyses if not accounted for [39, 40].

It is noteworthy that our results are consistent with previously published data. For example, in the meta-analysis by Peng et al. the sensitivity of digital PCR methods to detect *KRAS* mutation in plasma samples of CRC patients ranged from 67% to 96% in 12 studies; however, the results varied greatly due to limited sample sizes and utilization of different digital PCR methods [41]. In another meta-analysis comprising 17 studies that used ddPCR to detect *KRAS* mutation in cfDNA, sensitivity ranged from 36% to 100% and specificity from 50 % to 100 % [42]. Bettegowda et al. reported that the sensitivity of ctDNA for detection of clinically relevant *KRAS* gene mutations was 87.2% and its specificity was 99.2% using digital PCR in 206 metastatic colorectal cancer patients [6]. The higher sensitivity in the study by Bettegowda et al. can be explained by the fact that only patients with metastatic disease were included. Accordingly, we attempted to analyze only cases of metastatic CRC and the sensitivity increased to 87.0%. Likewise, a study focused on ctDNA analysis in patients divided according to tumor stage confirmed that ctDNA concentrations increase with tumor size and cancer stage. For example, CRC patients in stage I showed 47% sensitivity of *KRAS* mutation in contrast to 87% sensitivity observed in patients in stage IV [43]. This assumption is also supported by a study by Liebs et al., who measured the mutations in matching tumor and plasma samples and observed the highest accuracy (68%) in patients with distant metastases, demonstrating that cfDNA analysis by ddPCR in patients with earlier stages of cancer is limited [44].

Interestingly, we observed that presence of any *KRAS, BRAF*, or *NRAS* mutation was more prevalent in right-sided colorectal cancers, particularly those originating in the cecum (25.4% vs. 8.5%, p = 0.009). Moreover, patients with these mutations had a higher frequency of advanced nodal involvement (N2: 41.3% vs. 23.4%, p = 0.049) and a greater proportion of stage IV disease (47.6% vs. 36.2%, p = 0.017), suggesting an association between mutational status and more aggressive tumor behavior. These findings are in agreement with previous studies demonstrating that *RAS* and *BRAF* mutations are more common in right-sided colorectal cancers and are associated with poorer prognosis [15, 45-48]. While our cohort was relatively small, the observed trends reinforce the clinical value of liquid biopsy in capturing actionable mutational patterns and guiding treatment decisions based on tumor location and molecular profile.

## 5. Conclusions

Liquid biopsy collection followed by ctDNA analysis represents a promising alternative to the standard care examination of tissue obtained by biopsy. Its use is particularly suggested for monitoring cancer relapse after primary treatment or as a tool to assess the response to treatment in patients with metastasis by monitoring minimal residual disease through the detection of specific tumor mutations in peripheral blood.

Our data support the potential of ddPCR being a reliable and sensitive method for the detection and analysis of ctDNA in molecular diagnosis of colorectal cancer. However, the routine use of this approach in the management of CRC patients remains to be widely applied in clinical practice. Herein lies the importance of our work, which demonstrates that liquid biopsy may represent a valuable complement to standard tissue-based biopsy in molecular diagnosis of CRC patients because it overcomes the problem of tumor heterogeneity and allows detection of mutations that may be missed due to spatial and temporal subsampling of tumor tissue or due to a minor number of mutated cells. Thus, we conclude that liquid biopsy and ctDNA analysis by ddPCR may represent a complementary tool to standard tissue-based biopsy.

## Acknowledgments

We would like to thank Tamara Kolarova for her excellent technical support.

## Data Availability Statement

The data presented in this study are available on request from the corresponding author. The data are not publicly available due to privacy restrictions.

## Authors’ Contributions

Conceptualization, T.K., I.K. and R.H.; methodology, J.B., J.H., K.M. I.K. and R.H.; validation, K.O., M.C. and M.U.; formal analysis, J.B. and M.B.; investigation, K.O., M.C., J.H., O.H. and K.M.; resources, J.B. and R.H.; data curation, M.C., M.U. and J.H:; writing—original draft preparation, K.O., M.C. and O.H.; writing—review and editing, J.H., M.B. T.K., K.M., I.K. and R.H.; visualization, K.O. and M.C.; supervision, T.K., I.K. and R.H.; project administration, J.H.; funding acquisition, R.H. All authors have read and agreed to the published version of the manuscript.

## Ethics approval and consent to participate

The study was conducted in accordance with the Declaration of Helsinki and approved by Ethics Committee of Masaryk Memorial Cancer Institute (protocol code 2020/2489/MOU, approved 15. 9. 2020). Informed consent was obtained from all subjects involved in the study.

## Patient consent for publication

Not applicable

