## Supplementary data for "Detection of *KRAS, NRAS* and *BRAF* mutations in liquid biopsy from patients with colorectal cancer"

**Table S1.** Cell line information

| **Cell line** | **Origin** | **Mutation** | | | **Medium** |
|---|---|---|---|---|---|
| | | *KRAS* | *NRAS* | *BRAF* | |
| **Caco-2** | Colon adenocarcinoma | wt | wt | wt | HG-DMEM |
| **HCT116** | Colon carcinoma | G13D/wt | wt | wt | RPMI1640 |
| **PANC-1** | Pancreatic ductal adenocar-cinoma | G12D/wt | *NA* | wt | HG-DMEM |
| **DLD-1** | Colon adenocarcinoma | G13D/wt | *NA* | wt | HG-DMEM |
| **MIA PaCa-2** | Pancreatic ductal adenocar-cinoma | G12C | *NA* | wt | HG-DMEM |
| **SW480** | Colon adenocarcinoma | G12V | *NA* | wt | HG-DMEM |
| **HT-29** | Colon adenocarcinoma | wt | wt | V600E/wt | HG-DMEM |
| **MOLT-4** | Adult T acute lymphoblastic leukemia | *NA* | G12C/wt | *NA* | HG-DMEM |

* wt – *wild type,* NA – mutation not analyzed

**Table S2.** ddPCR protocol (Bio-Rad)

<table>
<tr><th>Cycling Step</th><th>Temperature, °C</th><th>Time</th><th>Ramp Rate</th><th>Cycles</th></tr>
<tr><td>Enzyme activation</td><td>95</td><td>10 min</td><td rowspan="4">~ 2°C/sec</td><td>1</td></tr>
<tr><td>Denaturation</td><td>94</td><td>30 sec</td><td rowspan="2">40</td></tr>
<tr><td>Annealing/extension</td><td>55</td><td>1 min</td></tr>
<tr><td>Enzyme deactivation</td><td>98</td><td>10 min</td><td>1</td></tr>
<tr><td>Hold</td><td>4</td><td>Infinite</td><td>~ 1°C/sec</td><td>1</td></tr>
</table>

**Table S3.** Determination of Limit of Detection (LoD) of ddPCR for *KRAS* mutation detection in serial dilutions of genomic DNA purified from DLD-1 cells.

| Concentration of gDNA (ng/μL) | Copies/μl |
|---|---|
| **11.429** | 7.965 ± 0.644 |
| **2.857** | 2.030 ± 0.231 |
| **0.714** | 0.516 ± 0.110 |
| **0.179** | 0.206 ± 0.096 |
| **0.045** | 0.045 ± 0.018 |
| **0.011** | 0.007 ± 0.016 |
| **0.003** | NA |

**Table S4.** Determination of mutations in a cohort of colorectal cancer (CRC) patients.

| ID | Conc. [ng/µl] | mut KRAS predictive pathology | mut NRAS predictive pathology | mut BRAF predictive pathology | KRAS Positive droplets (MUT)[1] | KRAS negative droplets (MUT)[2] | KRAS All droplets | MUT c | WT c | NRAS Positive droplets (MUT) | NRAS negative droplets (MUT) | NRAS All drop-lets | MUT c | WT c | BRAF Positive droplets (MUT) | BRAF negative droplets (MUT) | All droplets | MUT c | WT c |
|---|---|---|---|---|---|---|---|---|---|---|---|---|---|---|---|---|---|---|---|
| 1 | 0.300 | undetected | undetected | undetected | 5 | 37398 | 37403 | 0.16 | 26.30 | | | | | | 0 | 49940 | 49940 | 0.00 | 23.70 |
| 2 | 0.572 | undetected | undetected | undetected | 0 | 42960 | 42960 | 0.00 | 43.10 | | | | | | 0 | 54728 | 54728 | 0.00 | 48.30 |
| 3 | 1.600 | undetected | undetected | undetected | 3 | 47707 | 47710 | 0.07 | 96.60 | | | | | | 0 | 52699 | 52699 | 0.00 | 145.30 |
| 4 | 1.640 | undetected | undetected | undetected | 2 | 51920 | 51922 | 0.05 | 105.70 | | | | | | 0 | 55456 | 55456 | 0.00 | 103.60 |
| 5 | 0.610 | undetected | mut G13R (59%) | undetected | 2 | 56945 | 56947 | 0.04 | 37.00 | 3 | 29760 | 29763 | 0.12 | 1.90 | 0 | 53309 | 53309 | 0.00 | 41.40 |
| 6 | 1.050 | undetected | undetected | undetected | 0 | 52856 | 52856 | 0.00 | 59.00 | | | | | | 0 | 52507 | 52507 | 0.00 | 77.40 |
| 7 | 0.722 | undetected | undetected | undetected | 0 | 53621 | 53621 | 0.00 | 45.90 | | | | | | 0 | 50154 | 50154 | 0.00 | 59.10 |
| 8 | 1.300 | mut G12D (15%) | undetected | undetected | 3 | 59235 | 59238 | 0.06 | 93.10 | | | | | | | | | | |
| 9 | 0.472 | undetected | undetected | undetected | 1 | 51534 | 51535 | 0.02 | 33.30 | | | | | | 0 | 53519 | 53519 | 0.00 | 37.50 |
| 11 | 1.100 | undetected | undetected | undetected | 2 | 55396 | 55398 | 0.04 | 76.60 | | | | | | 0 | 51001 | 51001 | 0.00 | 87.10 |
| 12 | 0.421 | undetected | undetected | undetected | 0 | 56844 | 56844 | 0.00 | 27.00 | | | | | | 0 | 55961 | 55961 | 0.00 | 22.30 |
| 13 | 0.691 | mut G13D (8%) | undetected | undetected | 2 | 54732 | 54734 | 0.04 | 57.40 | | | | | | | | | | |
| 14 | 0.744 | undetected | mut Q61R (30%) | undetected | 1 | 56068 | 56069 | 0.02 | 49.60 | 0 | 29488 | 29488 | 0.00 | 10.50 | 0 | 50628 | 50628 | 0.00 | 19.10 |
| 15 | 0.230 | mut G12D (21%) | undetected | undetected | 35 | 43584 | 43619 | 0.94 | 19.30 | | | | | | | | | | |
| 16 | 0.312 | mut G12D (23%) | undetected | not tested | 3 | 50754 | 50757 | 0.07 | 36.20 | | | | | | | | | | |

| | | | | | | | | | | | | | | | | | | | |
|---|---|---|---|---|---|---|---|---|---|---|---|---|---|---|---|---|---|---|---|
| 17 | 0.730 | undetected | undetected | undetected | 1 | 50963 | 50964 | 0.02 | 92.60 | | | | | | 0 | 37898 | 37898 | 0.00 | 14.90 |
| 18 | 2.960 | undetected | undetected | mut V600E (34%) | 5 | 60610 | 60615 | 0.10 | 397.00 | | | | | | 5 | 40303 | 40308 | 0.15 | 114.70 |
| 19 | 0.807 | mut G12V (34%) | undetected | undetected | 5 | 58744 | 58749 | 0.10 | 83.90 | | | | | | | | | | |
| 20 | 0.699 | mut G13D (30%) | undetected | undetected | 5 | 58021 | 58026 | 0.10 | 111.50 | | | | | | | | | | |
| 21 | 0.491 | mut G12D (30%) | undetected | undetected | 7 | 55852 | 55859 | 0.15 | 41.50 | | | | | | | | | | |
| 22 | 1.170 | undetected | undetected | mut V600E (19%) | 3 | 39073 | 39076 | 0.09 | 79.50 | | | | | | 2 | 39267 | 39269 | 0.06 | 54.40 |
| 23 | 1.290 | undetected | mut G13V (29%) | undetected | 1 | 61433 | 61434 | 0.02 | 99.10 | 0 | 32101 | 32101 | 0.00 | 8.60 | | | | | |
| 24 | 1.160 | undetected | undetected | mut V600E (15%) | 1 | 61274 | 61275 | 0.02 | 100.40 | | | | | | 0 | 48968 | 48968 | 0.00 | 99.40 |
| 25 | 0.489 | mut G12F (38%) | undetected | undetected | 0 | 41113 | 41113 | 0.00 | 41.70 | | | | | | | | | | |
| 26 | 0.410 | undetected | undetected | undetected | 0 | 61555 | 61555 | 0.00 | 26.60 | | | | | | 0 | 46302 | 46302 | 0.00 | 5.10 |
| 27 | 0.587 | mut G12V (31%) | undetected | undetected | 268 | 59666 | 59934 | 5.30 | 46.10 | | | | | | | | | | |
| 28 | 0.541 | mut G13D (13% | undetected | undetected | 2 | 59313 | 59315 | 0.04 | 64.20 | | | | | | | | | | |
| 29 | 0.150 | mut G12D (28%) | undetected | undetected | 18 | 50383 | 50401 | 0.42 | 12.60 | | | | | | | | | | |
| 30 | 0.451 | mut G13D (29%) | undetected | undetected | 2 | 37264 | 37266 | 0.06 | 35.40 | | | | | | | | | | |
| 32 | 0.954 | undetected | undetected | undetected | 3 | 59723 | 59726 | 0.06 | 131.00 | | | | | | 0 | 49600 | 49600 | 0.00 | 106.40 |

| | | | | | | | | | | | | | | | | | | | |
|---|---|---|---|---|---|---|---|---|---|---|---|---|---|---|---|---|---|---|---|
| 33 | 1.560 | mut G12S (57%) | undetected | undetected | 3 | 41444 | 41447 | 0.09 | 58.80 | | | | | | | | | | |
| 34 | 1.960 | mut G13D (40%) | undetected | undetected | 1026 | 61883 | 62909 | 19.30 | 129.90 | | | | | | | | | | |
| 35 | 0.339 | undetected | undetected | undetected | 1 | 59494 | 59495 | 0.02 | 24.50 | | | | | | 0 | 48133 | 48133 | 0.00 | 24.50 |
| 38 | 0.370 | mut G12D | undetected | undetected | 1 | 38422 | 38423 | 0.03 | 23.30 | | | | | | | | | | |
| 39 | 1.320 | mut G13D (43%) | undetected | undetected | 644 | 34640 | 35284 | 21.70 | 68.50 | | | | | | | | | | |
| 40 | 0.445 | undetected | mut Q61R (78%) | undetected | 1 | 59291 | 59292 | 0.02 | 28.90 | 0 | 39779 | 39779 | 0.00 | 1.18 | 0 | 49355 | 49355 | 0.00 | 14.50 |
| 41 | 0.481 | undetected | undetected | undetected | 0 | 57531 | 57531 | 0.00 | 29.90 | | | | | | 0 | 53315 | 53315 | 0.00 | 36.00 |
| 42 | 0.184 | mut G13C (22%) | undetected | undetected | 0 | 37946 | 37946 | 0.00 | 10.80 | | | | | | | | | | |
| 43 | 1.420 | undetected | undetected | mut V600E (18%) | 4 | 51962 | 51966 | 0.09 | 107.50 | | | | | | 5 | 50216 | 50221 | 0.12 | 64.10 |
| 44 | 0.991 | undetected | undetected | mut V600E (29%) | 0 | 54668 | 54668 | 0.00 | 68.40 | | | | | | 0 | 50380 | 50380 | 0.00 | 72.90 |
| 45 | 0.383 | undetected | undetected | undetected | 1 | 56079 | 56080 | 0.02 | 27.10 | | | | | | 0 | 48166 | 48166 | 0.00 | 14.00 |
| 46 | 0.499 | undetected | undetected | mut V600E (22%) | 2 | 55811 | 55813 | 0.04 | 38.00 | | | | | | 329 | 53315 | 53644 | 1.62 | 7.20 |
| 47 | 0.371 | undetected | undetected | undetected | 0 | 55728 | 55728 | 0.00 | 27.70 | 0 | 31684 | 31684 | 0.00 | 30.50 | 0 | 53093 | 53093 | 0.00 | 23.00 |
| 48 | 0.847 | mut G12D (22%) | undetected | undetected | 255 | 55465 | 55720 | 5.40 | 53.70 | | | | | | | | | | |
| 49 | 0.842 | undetected | undetected | mut V600E(28%) | 3 | 59603 | 59606 | 0.06 | 64.70 | | | | | | 4 | 49995 | 49999 | 0.09 | 67.00 |
| 50 | 0.242 | undetected | undetected | mut V600E | 5 | 45700 | 45705 | 0.13 | 40.60 | | | | | | | | | | |
| 52 | 0.773 | undetected | undetected | undetected | 1 | 50283 | 50284 | 0.02 | 3.80 | 1 | 39732 | 39733 | 0.03 | 1.78 | | | | | |

| | | | | | | | | | | | | | | | | | | | |
|---|---|---|---|---|---|---|---|---|---|---|---|---|---|---|---|---|---|---|---|
| 53 | 0.230 | undetected | undetected | undetected | 3 | 57701 | 57704 | 0.06 | 57.10 | 0 | 49857 | 49857 | 0.00 | 25.20 | 0 | 49443 | 49443 | 0.00 | 57.10 |
| 54 | 1.280 | undetected | undetected | undetected | 3 | 58938 | 58941 | 0.06 | 15.50 | 0 | 47118 | 47118 | 0.00 | 0.37 | 0 | 51605 | 51605 | 0.00 | 13.40 |
| 55 | 1.270 | undetected | undetected | undetected | 1 | 30355 | 30356 | 0.04 | 95.50 | 0 | 50010 | 50010 | 0.00 | 32.10 | 0 | 53396 | 53396 | 0.00 | 93.40 |
| 56 | 2.580 | undetected | undetected | undetected | 1 | 52813 | 52814 | 0.02 | 0.00 | 0 | 47853 | 47853 | 0.00 | 33.80 | 0 | 53025 | 53025 | 0.00 | 139.00 |
| 57 | 33.900 | mut G12C | not tested | not tested | 5 | 49089 | 49094 | 0.12 | 208.00 | 16 | 47165 | 47181 | 0.40 | 1030.00 | | | | | |
| 58 | 9.690 | undetected | undetected | undetected | 31614 | 23049 | 54663 | 1016.00 | 1080.00 | | | | | | 0 | 50285 | 50285 | 0.00 | 76.10 |
| 59 | 11.400 | mut G12V (25%) | undetected | undetected | 10 | 59773 | 59783 | 0.20 | 217.70 | | | | | | | | | | |
| 60 | 1.620 | mut G13C (30%) | undetected | undetected | 7 | 39260 | 39267 | 0.21 | 828.00 | | | | | | | | | | |
| 61 | 0.876 | undetected | undetected | mut V600E (20%) | 5 | 59190 | 59195 | 0.10 | 113.30 | | | | | | 0 | 52941 | 52941 | 0.00 | 50.70 |
| 62 | 0.440 | undetected | undetected | undetected | 1 | 59336 | 59337 | 0.02 | 63.50 | | | | | | 0 | 50375 | 50375 | 0.00 | 37.40 |
| 63 | 0.692 | mut G12V (38%) | undetected | undetected | 1 | 39078 | 39079 | 0.03 | 32.60 | 3 | 50761 | 50764 | 0.07 | 24.40 | | | | | |
| 64 | 0.218 | undetected | undetected | undetected | 0 | 55845 | 55845 | 0.00 | 0.34 | | | | | | | | | | |
| 65 | 0.702 | mut G12D (30%) | undetected | undetected | 0 | 56589 | 56589 | 0.00 | 11.90 | 1 | 49483 | 49484 | 0.02 | 25.30 | | | | | |
| 67 | 0.148 | mut A146T | undetected | undetected | 0 | 40862 | 40862 | 0.00 | 0.63 | | | | | | | | | | |
| 69 | 0.231 | undetected | undetected | undetected | 4 | 55287 | 55291 | 0.09 | 13.30 | | | | | | | | | | |
| 70 | 0.164 | mut G12A | undetected | undetected | 0 | 54719 | 54719 | 0.00 | 0.15 | | | | | | | | | | |
| 72 | 0.118 | mut A146T | undetected | undetected | 5 | 54853 | 54858 | 0.11 | 9.90 | | | | | | | | | | |
| 73 | 1.400 | mut Q61/E62 | undetected | undetected | 13 | 56074 | 56087 | 0.27 | 111.90 | | | | | | | | | | |
| 74 | 1.610 | mut G12V | undetected | undetected | 12 | 57713 | 57725 | 0.24 | 81.10 | | | | | | | | | | |
| 75 | 0.105 | mut G12A | undetected | undetected | 1 | 58556 | 58557 | 0.02 | 3.06 | | | | | | | | | | |
| 76 | 0.252 | undetected | undetected | undetected | 6 | 57286 | 57292 | 0.12 | 11.10 | | | | | | | | | | |

| | | | | | | | | | |
|---|---|---|---|---|---|---|---|---|---|
| 77 | 0.161 | undetected | undetected | undetected | 23 | 56263 | 56286 | 0.48 | 19.80 |
| 78 | 0.114 | undetected | undetected | undetected | 46 | 56490 | 56536 | 0.96 | 4.50 |
| 79 | 0.253 | undetected | undetected | undetected | 7 | 54157 | 54164 | 0.15 | 20.60 |
| 80 | 0.625 | mut G12A | undetected | undetected | 15 | 56146 | 56161 | 0.31 | 36.40 |
| 81 | 1.530 | undetected | undetected | undetected | 13 | 56671 | 56684 | 0.27 | 119.80 |
| 82 | 0.630 | mut G12V | undetected | undetected | 9 | 52008 | 52017 | 0.20 | 40.80 |
| 83 | 0.646 | undetected | undetected | undetected | 20 | 57099 | 57119 | 0.41 | 52.50 |
| 84 | 0.553 | undetected | undetected | undetected | 14 | 53020 | 53034 | 0.31 | 36.20 |
| 85 | 0.145 | mut G12V | undetected | undetected | 21 | 48126 | 48147 | 0.51 | 7.80 |
| 86 | 0.673 | mut A146T | undetected | undetected | 4 | 57510 | 57514 | 0.08 | 36.10 |
| 87 | 2.640 | undetected | undetected | undetected | 19 | 51692 | 51711 | 0.43 | 170.00 |
| 88 | 0.468 | mut G12V | undetected | undetected | 18 | 56115 | 56133 | 0.38 | 28.70 |
| 89 | 0.837 | mut G12C | undetected | undetected | 20 | 52456 | 52476 | 0.45 | 68.90 |
| 90 | 0.415 | undetected | undetected | undetected | 0 | 56573 | 56573 | 0.00 | 0.19 |
| 91 | 2.500 | undetected | undetected | undetected | 14 | 53180 | 53194 | 0.31 | 144.20 |
| 94 | 0.310 | mut G12D | undetected | undetected | 1 | 54237 | 54238 | 0.02 | 16.00 |
| 95 | 0.113 | mut G13D | undetected | undetected | 0 | 50674 | 50674 | 0.00 | 5.20 |
| 97 | 0.672 | mut G12D | undetected | undetected | 1 | 55005 | 55006 | 0.02 | 38.20 |
| 98 | 0.935 | mut G12D | undetected | undetected | 2 | 53245 | 53247 | 0.04 | 42.80 |
| 99 | 0.730 | mut G12D | undetected | undetected | 5 | 58019 | 58024 | 0.10 | 29.30 |
| 100 | 0.369 | mut G12D | undetected | undetected | 25 | 62266 | 62291 | 0.47 | 9.90 |
| 101 | 7.410 | mut G12V | undetected | undetected | 5080 | 56380 | 61460 | 101.50 | 173.10 |
| 104 | 0.590 | undetected | undetected | undetected | 0 | 59701 | 59701 | 0.00 | 31.40 |
| 109 | 0.477 | mut G12S | undetected | undetected | 153 | 56638 | 56791 | 3.20 | 25.50 |
| 110 | 0.466 | undetected | undetected | undetected | 0 | 57731 | 57731 | 0.00 | 25.10 |
| 111 | 0.354 | mut G12C | undetected | undetected | 0 | 51801 | 51801 | 0.00 | 15.50 |
| 112 | 1.990 | mut G12V | undetected | undetected | 3 | 56261 | 56264 | 0.06 | 145.20 |

| | | | | | | | | | |
|---|---|---|---|---|---|---|---|---|---|
| 113 | 0.515 | undetected | undetected | undetected | 0 | 59179 | 59179 | 0.00 | 22.80 |
| 116 | 2.210 | mut G12S | undetected | undetected | 11 | 55384 | 55395 | 0.23 | 175.20 |
| 117 | 0.659 | undetected | undetected | undetected | 0 | 58275 | 58275 | 0.00 | 31.10 |
| 118 | 1.650 | undetected | undetected | undetected | 12 | 59918 | 59930 | 0.28 | 104.30 |
| 119 | 0.415 | undetected | undetected | undetected | 0 | 62630 | 62630 | 0.00 | 4.20 |
| 120 | 0.609 | undetected | undetected | undetected | 13 | 60954 | 60967 | 0.25 | 37.40 |
| 121 | 1.130 | undetected | undetected | undetected | 7 | 54513 | 54520 | 0.15 | 74.10 |
| 122 | 0.278 | undetected | undetected | undetected | 0 | 59264 | 59264 | 0.00 | 12.80 |
| 124 | 0.328 | undetected | undetected | undetected | 8 | 59765 | 59773 | 0.12 | 11.20 |
| 125 | 0.194 | undetected | undetected | undetected | 10 | 60070 | 60080 | 0.21 | 14.40 |
| 126 | 0.619 | mut G12C | undetected | undetected | 4 | 58360 | 58364 | 0.08 | 31.60 |
| 127 | 1.200 | undetected | undetected | undetected | 0 | 60856 | 60856 | 0.00 | 72.20 |
| 128 | 0.274 | mut G12C | undetected | undetected | 1 | 60653 | 60654 | 0.02 | 9.90 |
| 129 | 1.200 | mut G13D | undetected | undetected | 0 | 59768 | 59768 | 0.00 | 65.70 |
| 130 | 0.106 | mut G12D | undetected | undetected | 5 | 57561 | 57566 | 0.10 | 6.40 |

[1, 2] Positive and negative droplets indicate the absolute droplet counts detected by ddPCR. According to assay validation (LoB/LoD criteria), samples with two or more positive droplets were considered positive.

**Table S5.** Comparison of baseline patient characteristics according to the concordance of ddPCR and NGS in *KRAS* mutation.

| **Basic characteristics of patients and primary tumor** (n = 110 patients) | | **Discordant in KRAS mutation** (n = 45) | **Concordant in KRAS mutation** (n = 65) | **p-value** |
|---|---|---|---|---|
| **Sex** | women | 13 (28.9 %) | 26 (40 %) | 0.320 |
| | men | 32 (71.1 %) | 39 (60 %) | |
| **Age at surgery** | [years] | 64.2 ± 10.7<br>65.3 (58.5; 72.1) | 64.8 ± 10.7<br>66.1 (56.7; 73.1) | 0.783 |
| **Localization of the primary tumor** | Cecum (C18.0, C18.1) | 8 (17.8 %) | 12 (18.5 %) | 0.869 |
| | colon (C18.2–C18.7, C18.9) | 13 (28.9 %) | 15 (23.1 %) | |
| | lesions extending beyond the colon (C18.8) | 3 (6.7 %) | 5 (7.7 %) | |
| | rectosigmoideal connection (C19) | 7 (15.6 %) | 15 (23.1 %) | |
| | rectum (C20) | 14 (31.1 %) | 18 (27.7 %) | |
| **Extent of tumor (T)** | T1 | 1 (2.2 %) | 0 (0.0 %) | 0.502 |
| | T2 | 2 (4.4 %) | 6 (9.2 %) | |
| | T3 | 32 (71.1 %) | 40 (61.5 %) | |
| | T4 | 9 (20 %) | 18 (27.7 %) | |
| | Tx | 1 (2.2 %) | 1 (1.5 %) | |
| **Metastases in the lymph nodes (N)** | N0 | 18 (40 %) | 19 (29.2 %) | 0.161 |
| | N1 | 16 (35.6 %) | 18 (27.7 %) | |
| | N2 | 10 (22.2 %) | 27 (41.5 %) | |
| | Nx | 1 (2.2 %) | 1 (1.5 %) | |
| **Distant metastases (M)** | M0 | 24 (53.3 %) | 33 (50.8 %) | 0.314 |
| | M1 | 17 (37.8 %) | 30 (46.2 %) | |
| | Mx | 4 (8.9 %) | 2 (3.1 %) | |
| **Lymphovascular space invasion (L)** | L0 | 27 (60 %) | 35 (53.8 %) | 0.304 |
| | L1 | 11 (24.4 %) | 24 (36.9 %) | |
| | Lx | 7 (15.6 %) | 6 (9.2 %) | |
| **Grade** | G1 | 15 (33.3 %) | 12 (18.5 %) | 0.184 |
| | G2 | 20 (44.4 %) | 40 (61.5 %) | |
| | G3 | 8 (17.8 %) | 10 (15.4 %) | |
| | G4 | 1 (2.2 %) | 0 (0.0 %) | |
| | Gx | 1 (2.2 %) | 3 (4.6 %) | |
| **Clinical stage** | stage I | 1 (2.2 %) | 2 (3.1 %) | 0.794 |
| | stage II | 10 (22.2 %) | 14 (21.5 %) | |
| | stage III | 17 (37.8 %) | 19 (29.2 %) | |
| | stage IV | 17 (37.8 %) | 30 (46.2 %) | |
| **Neoadjuvant therapy** | no | 33 (73.3 %) | 60 (92.3 %) | **0.015** |
| | yes | 12 (26.7 %) | 5 (7.7 %) | |
| | Same day | 17 (37.8 %) | 36 (55.4 %) | 0.109 |

| | | | |
|---|---|---|---|
| **Time between blood and tissue sampling** | Within 7 days | 19 (42.2 %) | 18 (27.7 %) |
| | Within 30 days | 4 (8.9 %) | 7 (10.8 %) |
| | Over 30 days | 5 (11.1 %) | 4 (6.2 %) |

*Mean ± standard deviation and median (interquartile range) are given for continuous variables, and absolute (relative) frequencies are given for categorical variables. P-values for statistical significance of differences between groups were calculated using the Pearson chi-square test (for nominal variables), the t-test (for continuous variables), and the Mann–Whitney test (for ordinal variables). If the assumptions for the Pearson chi-square test or the t-test were not met, Fisher's exact test or the Mann–Whitney test was used instead.*

**Table S6.** Comparison of baseline patient characteristics according to the concordance of ddPCR and NGS in *KRAS/BRAF/NRAS* mutation.

| **Basic characteristics of patients and primary tumor** (n = 110 patients) | | **Discordant in KRAS/BRAF/NRAS mutation** (n = 41) | **Concordant in KRAS/BRAF/NRAS mutation** (n = 69) | **p-value** |
|---|---|---|---|---|
| **Sex** | women | 9 (22 %) | 30 (43.5 %) | **0.038** |
| | men | 32 (78 %) | 39 (56.5 %) | |
| **Age at surgery** | [years] | 63.7 ± 10.6<br>64.9 (58.5; 70.0) | 65.0 ± 10.7<br>66.1 (56.7; 73.2) | 0.539 |
| **Localization of the primary tumor** | Cecum (C18.0, C18.1) | 6 (14.6 %) | 14 (20.3 %) | 0.687 |
| | colon (C18.2–C18.7, C18.9) | 10 (24.4 %) | 18 (26.1 %) | |
| | lesions extending beyond the colon (C18.8) | 2 (4.9 %) | 6 (8.7 %) | |
| | rectosigmoideal connection (C19) | 8 (19.5 %) | 14 (20.3 %) | |
| | rectum (C20) | 15 (36.6 %) | 17 (24.6 %) | |
| **Extent of tumor (T)** | T1 | 1 (2.4 %) | 0 (0.0 %) | 0.309 |
| | T2 | 2 (4.9 %) | 6 (8.7 %) | |
| | T3 | 30 (73.2 %) | 42 (60.9 %) | |
| | T4 | 7 (17.1 %) | 20 (29 %) | |
| | Tx | 1 (2.4 %) | 1 (1.4 %) | |
| **Metastases in the lymph nodes (N)** | N0 | 18 (43.9 %) | 19 (27.5 %) | **0.006** |
| | N1 | 16 (39 %) | 18 (26.1 %) | |
| | N2 | 6 (14.6 %) | 31 (44.9 %) | |
| | Nx | 1 (2.4 %) | 1 (1.4 %) | |
| **Distant metastases (M)** | M0 | 24 (58.5 %) | 33 (47.8 %) | 0.524 |
| | M1 | 15 (36.6 %) | 32 (46.4 %) | |
| | Mx | 2 (4.9 %) | 4 (5.8 %) | |
| | L0 | 24 (58.5 %) | 38 (55.1 %) | 0.070 |

| | | | | |
|---|---|---|---|---|
| **Lymphovascular space invasion (L)** | L1 | 9 (22 %) | 26 (37.7 %) | |
| | Lx | 8 (19.5 %) | 5 (7.2 %) | |
| **Grade** | G1 | 15 (36.6 %) | 12 (17.4 %) | 0.063 |
| | G2 | 17 (41.5 %) | 43 (62.3 %) | |
| | G3 | 6 (14.6 %) | 12 (17.4 %) | |
| | G4 | 1 (2.4 %) | 0 (0.0 %) | |
| | Gx | 2 (4.9 %) | 2 (2.9 %) | |
| **Clinical stage** | stage I | 1 (2.4 %) | 2 (2.9 %) | 0.802 |
| | stage II | 10 (24.4 %) | 14 (20.3 %) | |
| | stage III | 15 (36.6 %) | 21 (30.4 %) | |
| | stage IV | 15 (36.6 %) | 32 (46.4 %) | |
| **Neoadjuvant therapy** | no | 30 (73.2 %) | 63 (91.3 %) | **0.023** |
| | yes | 11 (26.8 %) | 6 (8.7 %) | |
| **Time between blood and tissue sampling** | Same day | 15 (36.6 %) | 38 (55.1 %) | 0.182 |
| | Within 7 days | 19 (46.3 %) | 18 (26.1 %) | |
| | Within 30 days | 4 (9.8 %) | 7 (10.1 %) | |
| | Over 30 days | 3 (7.3 %) | 6 (8.7 %) | |

*Mean ± standard deviation and median (interquartile range) are given for continuous variables, and absolute (relative) frequencies are given for categorical variables. P-values for statistical significance of differences between groups were calculated using the Pearson chi-square test (for nominal variables), the t-test (for continuous variables), and the Mann–Whitney test (for ordinal variables). If the assumptions for the Pearson chi-square test or the t-test were not met, Fisher's exact test or the Mann–Whitney test was used instead.*

**Table S7.** Evaluation of the diagnostic accuracy of *KRAS* or any *KRAS/BRAF/NRAS* mutation detection by ddPCR – subgroup analysis.

| | **KRAS mutation** | **KRAS/BRAF/NRAS mutation** |
|---|---|---|
| **Clinical stage I+II** (n = 27) | | |
| **Area under the curve (AUC)** | 0.611 (0.410; 0.813) | 0.611 (0.410; 0.813) |
| **Sensitivity** | 66.7 (33.3; 88.9) % | 66.7 (33.3; 88.9) % |
| **Specificity** | 55.6 (33.0; 76.0) % | 55.6 (33.0; 76.0) % |
| **Positive predictive value** | 42.9 (20.6; 68.4) % | 42.9 (20.6; 68.4) % |
| **Negative predictive value** | 76.9 (47.8; 92.4) % | 76.9 (47.8; 92.4) % |
| **Clinical stage III** (n = 36) | | |
| **Area under the curve (AUC)** | 0.528 (0.360; 0.695) | 0.583 (0.406; 0.760) |
| **Sensitivity** | 55.6 (33.0; 76.0) % | 58.3 (38.3; 75.9) % |

| | | |
|---|---|---|
| **Specificity** | 50.0 (28.4; 71.6) % | 58.3 (30.8; 81.5) % |
| **Positive predictive value** | 52.6 (31.1; 73.2) % | 73.7 (50.2; 88.6) % |
| **Negative predictive value** | 52.9 (30.3; 74.5) % | 41.2 (21.0; 64.8) % |
| **Clinical stage IV** (n = 47) | | |
| **Area under the curve (AUC)** | 0.643 (0.520; 0.766) | 0.623 (0.484; 0.761) |
| **Sensitivity** | 87.0 (66.5; 95.7) % | 83.3 (65.7; 92.9) % |
| **Specificity** | 41.7 (24.1; 61.7) % | 41.2 (21.0; 64.8) % |
| **Positive predictive value** | 58.8 (41.9; 73.9) % | 71.4 (54.6; 83.9) % |
| **Negative predictive value** | 76.9 (47.8; 92.4) % | 58.3 (30.8; 81.5) % |
| **Without neoadjuvant therapy** (n = 93) | | |
| **Area under the curve (AUC)** | 0.658 (0.564; 0.751) | 0.665 (0.569; 0.761) |
| **Sensitivity** | 78.6 (63.7; 88.5) % | 76.9 (63.6; 86.4) % |
| **Specificity** | 52.9 (39.4; 66.1) % | 56.1 (40.8; 70.3) % |
| **Positive predictive value** | 57.9 (44.8; 69.9) % | 69.0 (56.0; 79.5) % |
| **Negative predictive value** | 75.0 (58.5; 86.4) % | 65.7 (48.8; 79.4) % |

For all values 95% CI is given.

**Table S8.** Comparison of baseline patient characteristics according to the presence of *KRAS* mutation by NGS.

| **Basic characteristics of patients and primary tumor** (n = 110 patients) | | **Without KRAS mutation by NGS** (n = 60) | **Presence of KRAS mutation by NGS** (n = 50) | **p-value** |
|---|---|---|---|---|
| **Sex** | women | 20 (33.3 %) | 19 (38.0 %) | 0.757 |
| | men | 40 (66.7 %) | 31 (62.0 %) | |
| **Age at surgery** | [years] | 64.4 ± 10.6<br>65.8 (59.6; 73.1) | 64.6 ± 10.9<br>66.0 (56.7; 72.2) | 0.907 |
| **Localization of the primary tumor** | Cecum (C18.0, C18.1) | 9 (15.0 %) | 11 (22.0 %) | 0.134 |
| | colon (C18.2–C18.7, C18.9) | 15 (25.0 %) | 13 (26.0 %) | |
| | lesions extending beyond the colon (C18.8) | 5 (8.3 %) | 3 (6.0 %) | |
| | rectosigmoideal connection (C19) | 17 (28.3 %) | 5 (10.0 %) | |
| | rectum (C20) | 14 (23.3 %) | 18 (36.0 %) | |
| **Extent of tumor (T)** | T1 | 0 (0.0 %) | 1 (2.0 %) | 0.587 |
| | T2 | 6 (10.0 %) | 2 (4.0 %) | |

| | | | | |
|---|---|---|---|---|
| | T3 | 40 (66.7 %) | 32 (64.0 %) | |
| | T4 | 13 (21.7 %) | 14 (28.0 %) | |
| | Tx | 1 (1.7 %) | 1 (2.0 %) | |
| **Metastases in the lymph nodes (N)** | N0 | 24 (40.0 %) | 13 (26.0 %) | 0.270 |
| | N1 | 19 (31.7 %) | 15 (30.0 %) | |
| | N2 | 16 (26.7 %) | 21 (42.0 %) | |
| | Nx | 1 (1.7 %) | 1 (2.0 %) | |
| **Distant metastases (M)** | M0 | 32 (53.3 %) | 25 (50.0 %) | 0.735 |
| | M1 | 24 (40.0 %) | 23 (46.0 %) | |
| | Mx | 4 (6.7 %) | 2 (4.0 %) | |
| **Lymphovascular space invasion (L)** | L0 | 33 (55.0 %) | 29 (58.0 %) | 0.659 |
| | L1 | 21 (35.0 %) | 14 (28.0 %) | |
| | Lx | 6 (10.0 %) | 7 (14.0 %) | |
| **Grade** | G1 | 15 (25.0 %) | 12 (24.0 %) | 0.986 |
| | G2 | 33 (55.0 %) | 27 (54.0 %) | |
| | G3 | 9 (15.0 %) | 9 (18.0 %) | |
| | G4 | 1 (1.7 %) | 0 (0.0 %) | |
| | Gx | 2 (3.3 %) | 2 (4.0 %) | |
| **Clinical stage** | stage I | 3 (5.0 %) | 0 (0.0 %) | 0.378 |
| | stage II | 15 (25.0 %) | 9 (18.0 %) | |
| | stage III | 18 (30.0 %) | 18 (36.0 %) | |
| | stage IV | 24 (40.0 %) | 23 (46.0 %) | |
| **Neoadjuvant therapy** | no | 51 (85.0 %) | 42 (84.0 %) | 1.000 |
| | yes | 9 (15.0 %) | 8 (16.0 %) | |

*Mean ± standard deviation and median (interquartile range) are given for continuous variables, and absolute (relative) frequencies are given for categorical variables. P-values for statistical significance of differences between groups were calculated using the Pearson chi-square test (for nominal variables), the t-test (for continuous variables), and the Mann–Whitney test (for ordinal variables). If the assumptions for the Pearson chi-square test or the t-test were not met, Fisher's exact test or the Mann–Whitney test was used instead.*

**Table S9.** Comparison of baseline patient characteristics according to the presence of any *KRAS*/*BRAF*/*NRAS* mutation by NGS.

| **Basic characteristics of patients and primary tumor** (n = 110 patients) | | **Without KRAS/BRAF/NRAS mutation by NGS** (n = 47) | **Presence of KRAS/BRAF/NRAS mutation by NGS** (n = 63) | **p-value** |
|---|---|---|---|---|
| **Sex** | women | 15 (31.9 %) | 24 (38.1 %) | 0.639 |
| | men | 32 (68.1 %) | 39 (61.9 %) | |
| **Age at surgery** | [years] | 62.9 ± 11.0<br>64.3 (57.9; 71.0) | 65.7 ± 10.4<br>67.3 (58.7; 74.5) | 0.226 |
| | Cecum (C18.0, C18.1) | 4 (8.5 %) | 16 (25.4 %) | **0.009** |

| | | | | |
|---|---|---|---|---|
| **Localization of the primary tumor** | colon (C18.2–C18.7, C18.9) | 12 (25.5 %) | 16 (25.4 %) | |
| | lesions extending beyond the colon (C18.8) | 4 (8.5 %) | 4 (6.3 %) | |
| | rectosigmoideal connection (C19) | 16 (34.0 %) | 6 (9.5 %) | |
| | rectum (C20) | 11 (23.4 %) | 21 (33.3 %) | |
| **Extent of tumor (T)** | T1 | 0 (0.0 %) | 1 (1.6 %) | 0.226 |
| | T2 | 6 (12.8 %) | 2 (3.2 %) | |
| | T3 | 31 (66.0 %) | 41 (65.1 %) | |
| | T4 | 9 (19.1 %) | 18 (28.6 %) | |
| | Tx | 1 (2.1 %) | 1 (1.6 %) | |
| **Metastases in the lymph nodes (N)** | N0 | 22 (46.8 %) | 15 (23.8 %) | **0.049** |
| | N1 | 13 (27.7 %) | 21 (33.3 %) | |
| | N2 | 11 (23.4 %) | 26 (41.3 %) | |
| | Nx | 1 (2.1 %) | 1 (1.6 %) | |
| **Distant metastases (M)** | M0 | 28 (59.6 %) | 29 (46.0 %) | 0.353 |
| | M1 | 17 (36.2 %) | 30 (47.6 %) | |
| | Mx | 2 (4.3 %) | 4 (6.3 %) | |
| **Lymphovascular space invasion (L)** | L0 | 29 (61.7 %) | 33 (52.4 %) | 0.620 |
| | L1 | 13 (27.7 %) | 22 (34.9 %) | |
| | Lx | 5 (10.6 %) | 8 (12.7 %) | |
| **Grade** | G1 | 15 (31.9 %) | 12 (19.0 %) | 0.233 |
| | G2 | 25 (53.2 %) | 35 (55.6 %) | |
| | G3 | 5 (10.6 %) | 13 (20.6 %) | |
| | G4 | 1 (2.1 %) | 0 (0.0 %) | |
| | Gx | 1 (2.1 %) | 3 (4.8 %) | |
| **Clinical stage** | stage I | 3 (6.4 %) | 0 (0.0 %) | **0.017** |
| | stage II | 15 (31.9 %) | 9 (14.3 %) | |
| | stage III | 12 (25.5 %) | 24 (38.1 %) | |
| | stage IV | 17 (36.2 %) | 30 (47.6 %) | |
| **Neoadjuvant therapy** | no | 41 (87.2 %) | 52 (82.5 %) | 0.684 |
| | yes | 6 (12.8 %) | 11 (17.5 %) | |

*Mean ± standard deviation and median (interquartile range) are given for continuous variables, and absolute (relative) frequencies are given for categorical variables. P-values for statistical significance of differences between groups were calculated using the Pearson chi-square test (for nominal variables), the t-test (for continuous variables), and the Mann–Whitney test (for ordinal variables). If the assumptions for the Pearson chi-square test or the t-test were not met, Fisher's exact test or the Mann–Whitney test was used instead.*
